# Communicative hand gestures as an implicit measure of artificial limb embodiment and daily usage

**DOI:** 10.1101/2020.03.11.20033928

**Authors:** Roni O. Maimon-Mor, Emeka Obasi, Jenny Lu, Nour Odeh, Stephen Kirker, Mairéad MacSweeney, Susan Goldin-Meadow, Tamar R. Makin

## Abstract

When people talk, they move their hands to enhance meaning. Here we ask whether people spontaneously use their artificial limbs (prostheses) to gesture, and whether prosthesis gesture behaviour relates to everyday prosthesis use and perceived embodiment. One-handed participants with congenital and acquired hand loss and two-handed controls participated in gesture-facilitating tasks, measured using acceleration monitors and further validated with offline video coding. Everyday functional prosthesis use and perceived prosthesis embodiment were assessed using questionnaires. Perhaps surprisingly, one- and two-handed participants did not differ in the amount of gestures they produced. However, they did differ in their gesture profile. One-handers performed more, and bigger, movements with their intact hand while gesturing relative to their prosthesis, whereas two-handers produced more equal movements across hands. Importantly, one-handers who incorporated their prosthesis more into gesturing, that is — produced gestures that were more similar to their two-handed counterparts — also showed more frequent prosthesis use in day-to-day life. Although as a group, one-handers only marginally agreed that their prosthesis feels like a body-part, people reporting positive embodiment also showed great prosthesis habits, both for communication and daily function. We propose that measuring gesture behaviour in prosthesis-users can be used as an implicit and objective clinical tool to monitor and assess successful prosthesis adoption.

## Introduction

Artificial limb technologies increasingly mirror the appearance and function of the human body (Vujaklija, Farina, & Aszmann, 2016). The hope is that greater similarity between an artificial limb and a natural hand will enable the users to perceive/relate to the prosthesis like a part of their own body, in other words, achieve ‘technological embodiment’ (Makin, de Vignemont, & Faisal, 2017). However, the relationship between successful prosthesis use and perceived prosthesis embodiment has not yet been demonstrated. Despite the technological progression, individuals with congenital and acquired missing upper-limb (hereafter one-handers) continue to report low functionality and use of their prostheses, and instead prefer to over-rely on their intact hand (Jang et al., 2011). In at least 30% of cases, one-handers abandon their prostheses altogether (Engdahl et al., 2015), often after being fitted with a customised prothesis (Østlie et al., 2012), resulting in wasted resources. Successful prosthesis use is difficult to predict, and often can only be determined by trial and error over the course of months. A further challenge is in quantifying how one-handers use their prostheses in day-to-day life, with research indicating that self-reports might be misleading (Pareés et al., 2012). Previous studies have overly focused on prosthesis dexterity (e.g., grasping and manipulating objects), which is not necessarily the best way to capture prosthesis adoption (See Discussion). Here, we focus on the significant role that our hands play in a core aspect of human life –– communication.

Gestures have an important communicative role. Not only do they increase listeners’ comprehension of speech, but they can also convey information that is not expressed in words (Goldin-Meadow, 2003; Goldin-Meadow & Alibali, 2013). Hand gestures are a ubiquitous aspect of upper limb use; they are observed across languages and cultures (Feyersein & De Lannoy, 1991), and even in congenitally blind individuals who have had no gesturing model to copy or learn from (Iverson & Goldin-Meadow, 1998). Gestures may therefore provide information about the extent to which individuals relate to their artificial limbs as hands. In this study, we aimed to characterise the use of upper-limb prostheses in gestures, using accelerometers, and investigate how prosthesis gesturing relates to prosthesis use during everyday life and perceived prosthesis embodiment. One-handers engaged in two tasks designed to probe gesticulation (Figure 1A) while being naïve to the purpose of the study. Daily prosthesis-use and perceived sense of embodiment of the prosthetic limb were measured using questionnaires. We hypothesised that better prosthesis use in daily activities will relate to more positive prosthesis embodiment and more prosthesis gesture.

**Figure 1.**
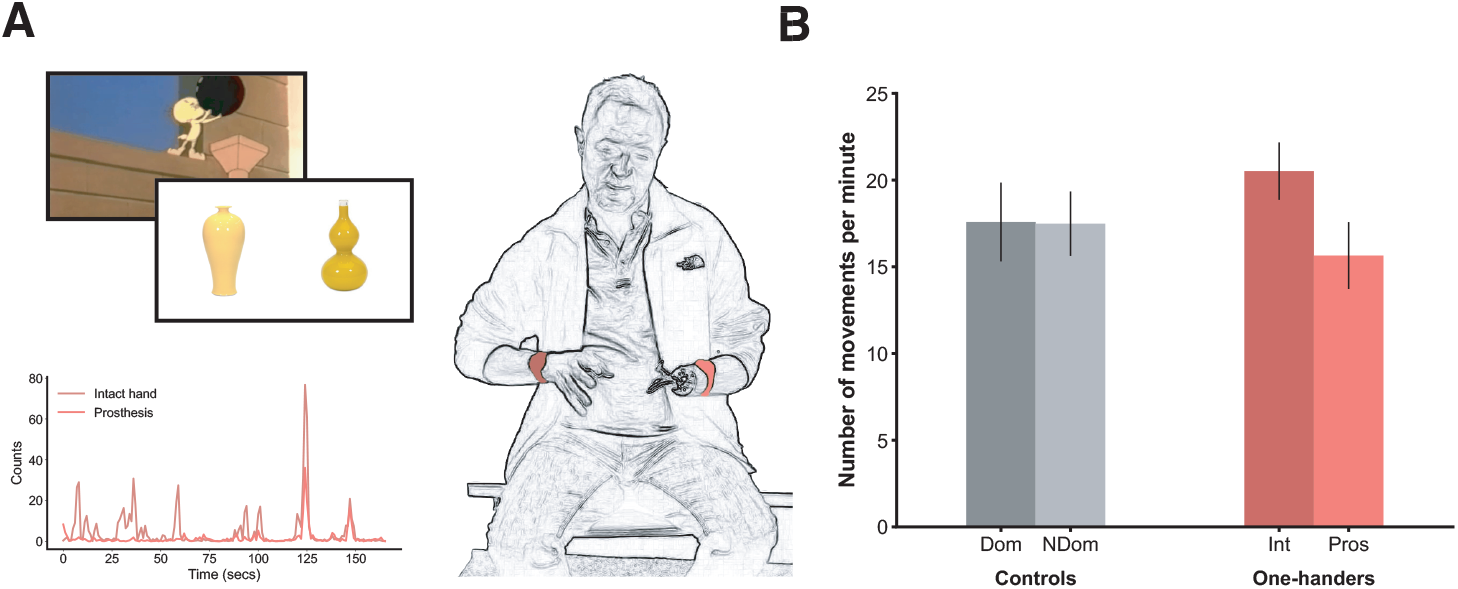
(A) *Experimental paradigm*. Top left: example stimuli from the paired object task and a frame from an animated video shown during the storytelling task. In the Paired Objects task, participants were asked to describe images of object pairs that looked very similar to one-another, specifically chosen to be difficult to convey using verbal description alone (Lu & Goldin-Meadow, 2018). In the storytelling task, participants watched two short animated video clips, then recalled and described them in as much detail as possible to a (presumed to be) naïve listener (McNeill, 1992; McNeill & Levy, 1982). Bottom left: pre-processed accelerometrydata of an example one-handed participant gesturing with his arms during the tasks. Dark red indicates measured acceleration of the intact hand, light red indicates prosthesis. Right: An illustration of a one-handed participant wearing the watch-like acceleration monitors used to measure gesticulation behaviour (B) *Number of movements analysis*. No significant difference between one-handers and two-handers in the amount of gestures performed per minute talking overall. However, we did find an interaction between arm and group (F_(1,38)_=4.25, p=0.046). One handers perform more movements with their intact arm compared to their prosthesis (t_(24)_= 2.94, p=0.007), while two-handers produced an equal amount of movements with both arms (t_(14)_= 0.088, p=0.93, BF_10_=0.263). Dom=Dominant arm, NDom=Nondominant arm, In=Intact arm, Pros= Prosthetic limb.

## Results

We first examined the relationship between daily prosthesis use and perceived prosthesis embodiment in our larger cohort of one-handed individuals (n=44; Tables 1 and S1). We found a significant correlation between these two measures (rho_(42)_ = 0.53, p < 0.001), revealing that prosthesis use and prosthesis embodiment are closely linked (Figure 2D). However, self-report questionnaire-based measures of embodiment are arguably crude and prone to bias and inter-individual differences (e.g. in interpretation of the written instructions). We therefore turned to a more implicit measure of embodiment––using the hand to gesture.

**Table 1.**
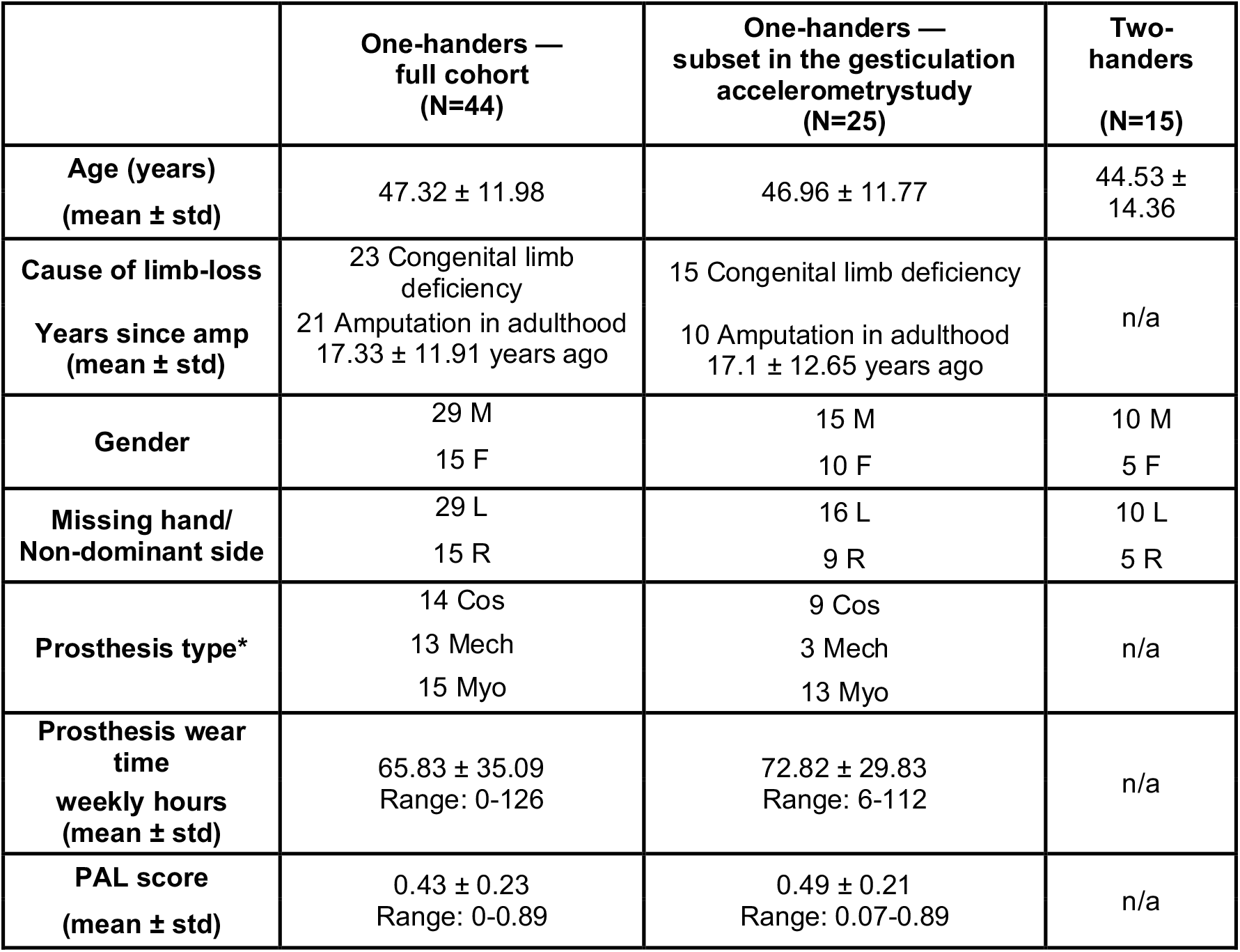
Demographic information on participants. Gender: M = male, F = female. Missing hand in one-handers and non-dominant hand in two-handers: R = right hand, L = left hand; Amp level = level of limb loss: Pros type = prosthesis type worn for the greatest time in a typical week: Cos = cosmetic, Mech = mechanical, Myo = myo-electric. Pros wear time = hours per week that prosthesis was typically worn. PAL score = functional ability with prosthesis as determined by PAL questionnaire: 0 = minimum function, 1 = maximum function. *Prosthesis type is not reported for 2 individuals who had a prosthesis they could wear but do not use it at all.

|  | One-handers —<br>full cohort<br>(N=44) | One-handers —<br>subset in the gesticulation<br>accelerometry study<br>(N=25) | Two-handers<br>(N=15) |
| --- | --- | --- | --- |
| **Age (years)**<br>(mean ± std) | 47.32 ± 11.98 | 46.96 ± 11.77 | 44.53 ± 14.36 |
| **Cause of limb-loss**<br>**Years since amp**<br>(mean ± std) | 23 Congenital limb deficiency<br>21 Amputation in adulthood<br>17.33 ± 11.91 years ago | 15 Congenital limb deficiency<br>10 Amputation in adulthood<br>17.1 ± 12.65 years ago | n/a |
| **Gender** | 29 M<br>15 F | 15 M<br>10 F | 10 M<br>5 F |
| **Missing hand/**<br>**Non-dominant side** | 29 L<br>15 R | 16 L<br>9 R | 10 L<br>5 R |
| **Prosthesis type\*** | 14 Cos<br>13 Mech<br>15 Myo | 9 Cos<br>3 Mech<br>13 Myo | n/a |
| **Prosthesis wear**<br>**time**<br>**weekly hours**<br>(mean ± std) | 65.83 ± 35.09<br>Range: 0-126 | 72.82 ± 29.83<br>Range: 6-112 | n/a |
| **PAL score**<br>(mean ± std) | 0.43 ± 0.23<br>Range: 0-0.89 | 0.49 ± 0.21<br>Range: 0.07-0.89 | n/a |

**Figure 2.**
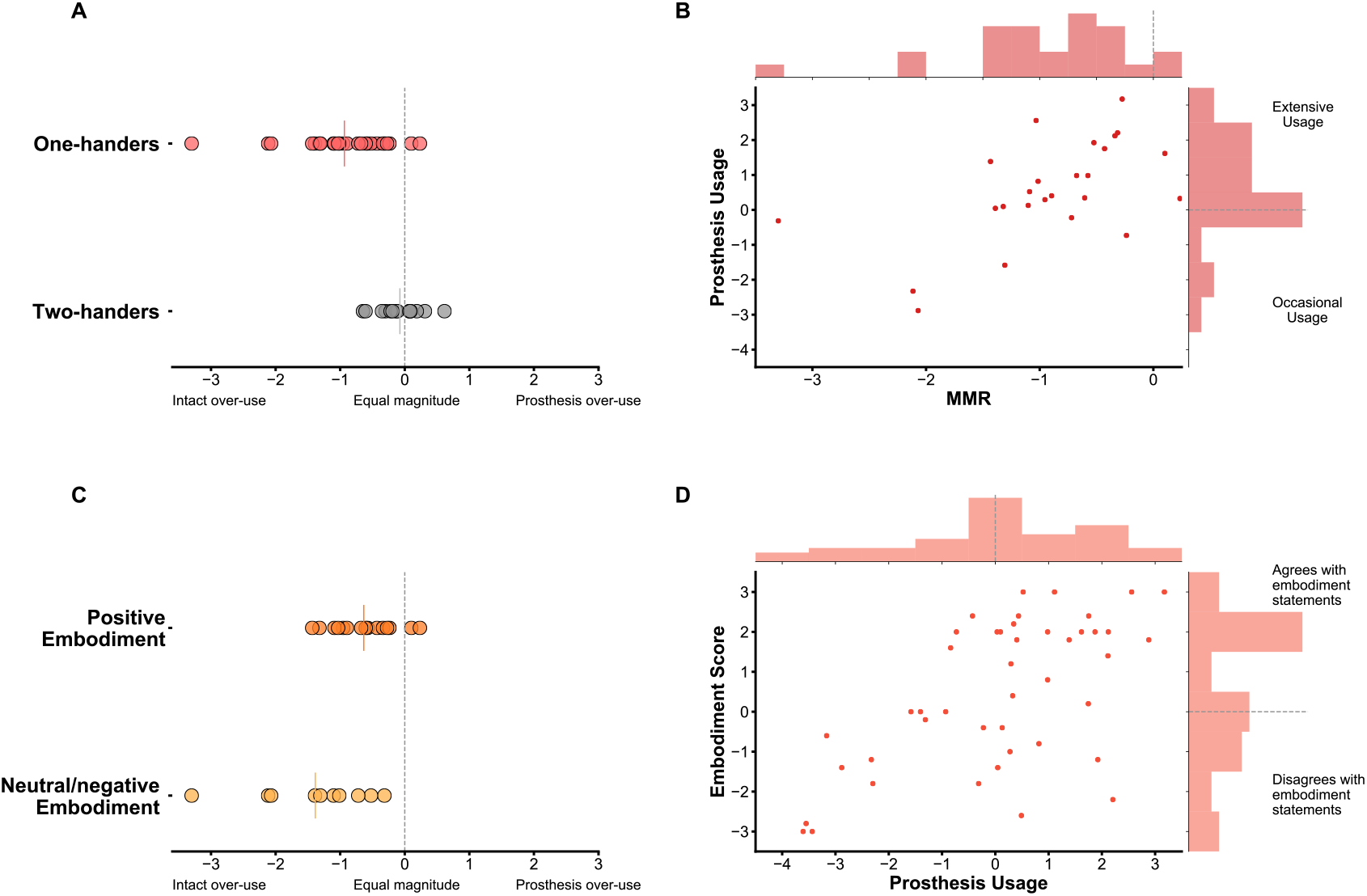
(A) MMR values across groups; two-handers performed relatively equal size arm movements when gesturing, whereas one-handers were significantly lateralised towards their intact arm (U = 47, p < 0.001). (B) Increased daily usage (measured by questionnaires) associated with increased incorporation of the prosthesis into gestures (measured by MMR) (rho_(23)_ = 0.55, p = 0.005) (C) MMR values across individuals who responded positively vs. neutral/negatively to prosthesis embodiment statements. Individuals who positively embody their prosthesis show increased incorporation of their prosthesis into gestures (U = 35, p = 0.03). (D) Greater prosthesis usage is associated with greater perceived prosthesis embodiment (rho_(42)_ = 0.53, p < 0.001). In A&C solid coloured lines indicates the group mean MMR. In B&D, the dashed lines in the histograms indicate the location of zero.

### Number and magnitude of gesture movements with the prosthesis

Looking first at overall gesture behaviour, we found that one-handers and two-handers did not differ in the overall number of movements per minute (as measured by accelerometry) that they produced with their two arms during the gesture tasks (rmANOVA group effect: F_(1,38)_=0.045 p=0.83; Figure 1B). However, we did find an interaction between arm and group (F_(1,38)_=4.25, p=0.046): While two-handers produced an equal amount of movements with both arms (t_(14)_= 0.088, p=0.93, BF_10_=0.263), one-handers produced a greater number of movements with their intact arm compared to their prosthetic arm (t_(24)_= 2.94, p=0.007).

We next calculated the Median-Magnitude Ratio (MMR) (Bailey, Klaesner, & Lang, 2015; Chadwell, Kenney, Thies, Galpin, & Head, 2016). This is a relative (laterality) measure, that reflects how much each arm contributed to the overall size of gesture movements on a second-by-second basis. Relative to the number of movements, this is a better validated measure which does not depend on an arbitrary threshold to separate movements. The MMR is therefore a more sensitive measure than gesture numbers, which we implemented in all further analysis. We found that one-handers made bigger movements with their intact arm than with their prosthesis arm (negative MMR) relative to controls, who showed similar sized movements across the two arms (U= 47, p < 0.001; Figure 2A). Similar results were also found using an alternative laterality measure of prosthesis use in gesture, one that is sensitive to the presence of movement in each second but not to the magnitude of the movement (see Figure S2A).

The laterality of gesture movement magnitude (MMR) correlated significantly with a laterality measure extracted from offline video coding (rho_(18)_ = 0.76, p < 0.001; Figure S1A; see Supplementary methods and Figure S1B for further validation). Thus, although one-handers gestured just as much as two-hander controls, the distribution of gestures across hands, and the character of these gestures, differed: one-handers performed more and larger movements with their intact arm than with their prosthesis arm; two-handers performed relatively symmetrical movements with their two arms.

Our one-handed sample consists of two sub-groups, individuals with congenital limb loss and amputees, who are known to adopt different adaptive strategies to compensate for their missing limb (Makin et al., 2013). We therefore determined whether cause of limb loss has an effect on prosthesis use during gesture. No significant effect was found when directly comparing the laterality in magnitude (MMR) or amount (numbers of movements) across the two arms of the two sub-groups (MMR: U=59, p=0.4; numbers of movements: U=66, p=0.64). Another way in which one-handers differ is the type of prosthesis they use. Nine of our participants used a cosmetic prosthesis, a passive hand-shaped apparatus; 16 used an active prosthesis, either a mechanical hook (n=3) or myoelectric prosthesis (n=13). Again, no significant effect was found when directly comparing between the laterality value of users of the two prosthesis types (cosmetic and active; MMR: U=71, p=0.98; number of movements: U=69, p=0.89). The type of prosthesis thus does not affect how it will be incorporated into co-speech gesture.

### Prosthesis gesture and daily prosthesis use

We next examined the relationship between use of the prosthesis to gesture and use of the prosthesis for daily activities. We found that prosthesis users who incorporated their prosthesis into their gestures in greater magnitude (resulting in a higher MMR) also tended to have a higher prosthesis use score, based on questionnaires probing functionality and wear frequency (rho_(23)_ = 0.55, p = 0.005; Figure 2B; see Figure S2B for similar results with the alternative measure). This effect is robust, and remains significant when controlling for cause of limb-loss and prosthesis type (see Supplementary results). Greater prosthesis use during daily life is thus associated with greater use of the prosthesis while gesturing.

We next examined this link between prosthesis use in gestures and use in daily life in relation to cause of amputation (congenital vs. acquired) and prosthesis type (passive vs. active) using parametric statistics. To meet the requisite statistical assumptions, we removed an outlier on the laterality measure (participant code:’aa11’) from the analysis. An ANCOVA with sub-group and prosthesis type as fixed-effects and daily prosthesis use score as a covariate revealed no significant sub-group effect on the laterality measure during gesture (cause of limb loss: F_(1,20)_=0.8, p=0.38; prosthesis type: F_(1,20)_<0.01, p=0.99). Cause of limb loss, or type of prosthesis, thus do not appear to play a key role in determining gesticulation behaviour with the prosthesis. Importantly, the relationship between prosthesis gesturing (as captured in the MMR) and daily prosthesis use remained significant in this analysis (F_(1,20)_=13.97, p=0.001), which highlights the robustness of the relationship between use of the prosthesis to gesture and use of the prosthesis for daily activities. This additional analysis further confirms that one-handers’ gesture behaviour is strongly related to their level of prosthesis use in daily activities and not to the cause of amputation or type of prosthesis used.

### Prosthesis gesture and perceived prosthesis embodiment

We next examined the relationship between gesture movements and prosthesis embodiment. Individuals varied in their responses to the subjective embodiment statements, producing a range from -3 (strongly disagree) to 3 (strongly agree) where 0 is a neutral response. As a whole, one-handed participants tended to marginally, although significantly, report that they experienced embodiment of their prosthesis (mean = +0.75, difference from zero: t_(24)_=2.23, p=0.035). We found similar effects when we looked at the full study cohort (i.e., all of the participants including those whose data were not included in the accelerometry part of the study); the full cohort showed a trend towards positive embodiment (n=44, mean = 0.47; difference from zero t_(43)_=1.685, p=0.099). We divided the one-handed participants into two groups based on whether they reported positive (score > 0) or neutral/negative (score ≤ 0) embodiment of their prosthesis. We then looked at the gesture laterality profiles, and found that one-handers who reported positive embodiment (n=15) used their prosthesis more when gesturing (i.e., higher MMR) than one-handers who reported neutral/negative embodiment (U_(23)_ = 35, p = 0.03; Figure 2C). When analysing the full range of embodiment scores, we found a trend towards a positive relationship between embodiment score and the laterality of gesture magnitude (MMR) (rho_(23)_ = 0.37, p = 0.07; Figure S3): the more positively one-handers respond to embodiment statements regarding their prosthesis, the more symmetrical their gestures are. Subjective measures of perceived embodiment, thus, only marginally associate with spontaneous gesture. Therefore, it appears that the implicit measure of gesture laterality better captures day-to-day prosthesis use.

## Discussion

Here we demonstrate that prosthetic limbs are regularly used to produce co-speech gestures, offering further demonstration of the ubiquity of gesture production in human communication. Despite hand loss (either congenitally or through amputation later in life), and independent of prosthesis type, one-handers gesture just as much as two-handers do. However, the profiles of the gestures produced by one- vs. two-handers differed. One-handers preferred to produce lateralised gestures, favouring their intact hand in amount and magnitude, while two-handers preferred to produce symmetrical gestures that were equally dispersed across both hands. Furthermore, one-handers who used their prosthesis more while gesturing, and produced gestures more similar to their two-handed counterparts, also tended to show more positive prosthesis embodiment and greater prosthesis use in day-to-day life. To our knowledge, this is also the first time that a strong relationship has been demonstrated between reported prosthesis embodiment and everyday prosthesis use.

Measuring prosthesis use is challenging both conceptually and practically. This challenge stems from the variety of tasks involving hand function in daily life and the complexity of defining what successful prosthesis use actually is: dexterity, wear time, satisfaction, or the elusive and ill-defined sense of embodiment? Practically, current assessment tools either involve self-report questionnaires, which can include large biases (Pareés et al., 2012); or they involve a dexterity task that is highly influenced by the participant’s motivation, prosthesis type, and the specific way in which individuals make use of their prosthesis in their daily lives. Here we provide a radically different hallmark for cognition of prosthesis use––how do you (spontaneously) employ your hands to convey meaning when talking? We believe that incorporating a prosthetic limb in gesture reflects a natural yet easily quantifiable level of immersion of the prosthesis in the user’s body and behaviour, which is one of the ultimate goals of any human-machine interfaces.

Based on our findings, we propose that accelerometery-based gesticulation analysis of one-handers could be used as a simple and objective clinical measure of prosthesis embodiment and every-day use. Assessing gesticulation is quick (up to 10 minutes), requires no training, can be used across prosthesis types, and provides a quantitative and objective measure (see https://osf.io/r3v8w/ for analysis code). As such, gesticulation may provide an ideal point-of-care clinical assessment for tracking the efficacy of upper-limb rehabilitation over time. Importantly, gesticulation can be measured implicitly, thus minimising user and clinician biases. As gesticulation with a prosthesis requires minimal skill, it could be measured in new users and tracked as they learn. Further research should determine whether gesticulation during fitting/early training can predict prosthesis adoption, which is a key issue in prosthesis rehabilitation.

## Methods

### Participants

44 one-handed individuals were recruited for this study: 21 unilateral acquired amputees (mean age ± std = 48.67 ± 12.9, 18 male, 12 with intact right hand), and 23 individuals with congenital unilateral upper-limb loss (age ± std = 46.09 ±11.22, 11 male, 17 with intact right hand; see Tables 1 and S1 for full demographic details). Nineteen individuals from the full set of participants were excluded from the gesticulation-accelerometry analysis for the following reasons: Issues with data storage (n=7); trans-humeral level limb-loss (n=4); did not produce any co-speech gestures (n=3); did not participate in gesture task (n=2); rated their typical weekly prosthesis use as 0 hours (n=2); aware of the purpose of the task before participating (n=1). A total of 25 participants (10 acquired amputees and 15-congenital one-handers) were included in the gesticulation-accelerometrystudy together with 15 age, gender, and handedness matched two-handed controls (see Table 1). All participants filled in the prosthesis-use and prosthesis embodiment questionnaires. There was no significant difference between one-handers and two-handers in age (t_(38)_ = 0.565, p = 0.58). There was also no significant difference between one- and two-handers in gender (Pearson chi-square = 0.000, p = 1) and handedness during the study (intact hand in one-handers and dominant hand in controls; Pearson chi-square = 0.03, p = 1).

Participants were recruited to the study based on the guidelines in our ethical approval UCL (REC: 9937/001) and in accordance with the declaration of Helsinki. The following inclusion criteria were taken into consideration during recruitment: (1) 18 to 70 years old, (2) MRI safe (for the purpose of other tasks conducted in the scanner), (3) no previous history of mental disorders, (4) regarding one-handers, owned at least one type of prosthesis during recruitment, (5) for acquired amputees, amputation occurred at least 6 months before recruitment. All participants gave full written informed consent for their participation, data storage, and filming.

### Tasks

Participants engaged in two tasks in which they were presented with a series of short video clips and images designed to probe gesticulation. The first was a storytelling task, which is a well-established gesture elicitation task (McNeill, 1992; McNeill & Levy, 1982) in which gestures are spontaneously produced during narrative discourse. Participants were shown two video clips of the cartoon ‘Tweety and Sylvester’ (see Figure 1A). After each clip, a listener, who the participants were informed was naïve to the videos, entered the room and sat opposite the participant. Each participant was then required to recall and describe the videos back to the listener in as much detail as possible.

The second task was the Paired Objects task. In each of the 4 trials, participants were presented with images of two items and asked to describe them in as much detail as possible to the listener. Each image displayed a pair of similar looking objects, specifically chosen to be difficult to convey using verbal description alone, therefore optimal for eliciting gestures (see Figure 1A). This method was developed by Lu & Goldin-Meadow in a study which focused on the depiction of shape and size in deaf participants (Lu & Goldin-Meadow, 2018). In addition to the listener present in the room, the participants were informed that an additional person will watch the video of their description and should be able to recognise the images based on the description. This instruction was added to emphasise the need for a thorough description. The listener was included as previous research suggests that individuals gesture more when there is a visible listener, compared to no listener or a listener hidden behind a screen (Alibali, Heath, & Myers, 2001). The stimuli were displayed on a computer screen using a Microsoft PowerPoint presentation, each pair was on a separate slide. When the participants indicated that they had finished describing the current pair, the experimenter pressed a button to move to the next trial.

In both parts, participants were naïve to the purpose of the task as being aware that the task was designed to elicit gesture could have interfered with their performance. Participants were seated to face the camera, which was used to record the task..

### Gesture measurements

To capture gesture behaviour, GENEActiv accelerometers (ActivinsightsLtd, Kimbolton, Cambridgeshire, UK) and AX3 accelerometers (Axivity, Newcastle upon Tyne, UK) were used. An accelerometer was placed on each of the participant’s arms, on both wrists for control participants, and on intact wrist and ‘prosthesis wrist’ for one-handed participants. The participants were not informed of the function of the accelerometers prior to the task to minimise any effect it may have had on performance. The accelerometers were set to record tri-axial data with a sampling frequency of 100Hz and range of ±8g, as well as the time stamp for each recorded signal. Raw acceleration data was extracted and pre-processed using MATLAB (version R2017a; Mathworks, Natick, MA, USA). The data from the left and right upper-limb/prosthesis were first synchronised with each other using the recorded time stamps to account for any minor sampling frequency errors between each device. The data was then plotted and visually inspected for any anomalous recordings, and the plot of the data was synchronised with the video clip of the task to ensure that the correct portions of data were analysed.

Movements along the 3 axes were combined 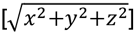 and bandpass filtered using a 4^th^ order Butterworth filter between the frequencies 0.2Hz and 15Hz to remove high frequency noise and gravitational artefact.

The filtered data was used to quantify gesture movements in two separate ways: (1) The total number of movements performed with each arm was calculated using a sliding window method, whereby an individual gesture was defined as each 400ms window of data in which there was movement (defined as an acceleration value ≥0.2g) that was preceded and succeeded by a window of no movement (<0.2g) (Makin et al., 2013). To account for differences in recording times between participants, the total number of movements performed per minute talking was calculated. (2) The median magnitude ratio (MMR) of the accelerometry data was calculated to investigate how much each arm contributed to the overall size of gesture movements performed during the task (Lang, Waddell, Klaesner, & Bland, 2017). This method has been previously used successfully to quantify every-day behaviour in impaired individuals and specifically amputees (Bailey, Klaesner, & Lang, 2015; Chadwell, Kenney, Thies, Galpin, & Head, 2016). Here, the data was down-sampled to 1Hz. The data was then converted into an arbitrary unit known as ‘counts’, where in each epoch, 1 count = 0.001664g of acceleration. The magnitude ratio (MR) between the intact arm and prosthesis was calculated for each second as 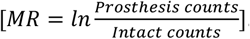. A value of 0 indicating equal movement of both arms, <0 indicating greater size movements with the intact/dominant arm relative to the prosthetic/non-dominant arm, and >0 indicating greater size movements with the prosthetic/non-dominant arm relative to the intact/dominant arm. To demonstrate that our results are not specific to a measure based on magnitude, the use ratio was also calculated. The use ratio quantifies the total duration of one arm’s movement with respect to the other (Lang et al., 2017). To calculate the use ratio, the number of seconds that included a substantial movement (a value larger than 12 count=∼0.02g) of the prosthetic/non-dominant arm was divided by the number of seconds that included a substantial movement of the intact/dominant arm. A value between 0 and 1 indicates greater use of the intact/dominant arm than the prosthetic/non-dominant arm; a value of 1 indicates equal use between both arms; and a value larger than 1 indicates greater use of the prosthetic/non-dominant arm than the intact/dominant arm.

Six participants (4 one-handers and 2 controls) only produced co-speech gestures in one of the tasks. For these participants, only data from the task during which they gestured was analysed; data from both tasks were analysed together for the remainder of the participants.

### Gesture measurements validation

To validate the accelerometry data, the movement laterality for a subset of the participants (n=20) was also calculated using offline video-coding of the Paired Objects task. Using the ELAN software, (ELAN v5.7, The Language Archive, Nijmegen, The Netherlands) (Lausberg & Slöetjes, 2008), separate gestures were manually coded and labelled based on their laterality. Gestures were labelled as follows: involving the dominant/intact hand only; involving the non-dominant/prosthesis hand only; or involving both hands/hand+prosthesis. The end of a gesture was identified based on a change in hand position, a change in verbal content, or by a return to resting position of the hands. For each participant and for each task, the percentage of gestures for each laterality label was calculated. A laterality index was then calculated as 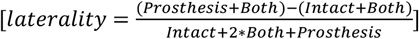, giving a value between -1 and +1, with 1 indicating total lateralisation towards the prosthetic/non-dominant hand, 0 indicating equal movement of both hands, and -1 indicating total lateralisation towards the intact/dominant hand. Coding reliability of the offline video-coding method was assessed by having an additional experimenter analyse a subset of 20 participants, and comparing the results between the two experimenters (see Supplementary figure S1B).

### Prosthesis Use Assessment

Participants completed a questionnaire to assess the frequency and functionality of prosthesis use, which were combined to create an overall prosthesis use score (as previously used in (Maimon-Mor & Makin, 2020; Van Den Heiligenberg et al., 2018; van den Heiligenberg, Yeung, Brugger, Culham, & Makin, 2017). To determine frequency of use, participants were asked to indicate the typical number of hours per day, and days per week, that they wear their prosthesis; these scores were then used to determine the typical number of hours per week that the prosthesis was worn. To determine functionality of prosthesis use, participants were asked to complete the prosthesis activity log (PAL) (Makin et al., 2013), a modified version of the Motor Activity Log (MAL) questionnaire, which is commonly used to assess arm functionality in those with upper-limb impairments (Uswatte, Taub, Morris, Light, & Thompson, 2006). The PAL consists of a list of 27 daily activities (see https://osf.io/jfme8/); participants must rate how often they incorporate their prosthesis to complete each activity on a scale of “never” (0 points), “sometimes” (1 point) or “very often” (2 points). The PAL score is then calculated as the participant’s score divided by the maximum possible score, generating a value between 0 (no functionality) and 1 (maximum functionality). Both prosthesis wear time and PAL were standardised using a Z-transform and summed to create a use score that included both wear time and incorporation of the prosthesis in activities of daily living. The two measurements (wear time and PAL) were highly correlated (Spearman’s rho=0.61, p=0.00001).

To validate the prosthesis usage questionnaire score, 21 participants completed the prosthesis use questionnaire twice, with 1-2 years between each measurement. Since the combined usage score is a sum of z-score transformation based on the specific dataset, we calculate the reliability of PAL and wear-time frequency separately. The PAL score was found to have excellent reliability with an ICC value of .81 (two-way random-model, absolute agreement type) and 95% confidence interval of single measures = .58 –.919 [F(20,20) = 10.60, p < .001]. For the wear time frequency, which is an ordinal 6-item non-symmetrical scale, we used Kendall’s tau-b correlation coefficient, showing a strong correlation in wear time scores (τ(19)=0.605, p=0.003). These analyses confirm that both measures have good consistency, and are a reliable measure for prosthesis use.

### Prosthesis Embodiment Assessment

The participants completed a 13-statement questionnaire to assess the extent of prosthesis embodiment (see https://osf.io/jfme8/). The statements were primarily adopted from a questionnaire used in rubber hand illusion studies, in which the embodiment of a rubber hand was investigated; “rubber hand” was replaced with “prosthesis” (Longo, Schüür, Kammers, Tsakiris, & Haggard, 2008). Questions were divided into the following factors: Body Ownership (embodiment), Agency, Body Image, and Somatosensory. The subset of embodiment statements used here are: “*it seems like the prosthesis belongs to me”, “it seems like the prosthesis is my hand”, “it seems like the prosthesis is part of my body”, “it feels like my prosthesis is a foreign body”, “it feels like my prosthesis is fused with my body”*. We did not analyse the results from the other control questions as we did not have a strong a priori hypothesis relating to these phenomena. We will make our full data available as an open source following publication. The participants rated each of these statements on a Likert scale from -3 (strongly disagree) to +3 (strongly agree). The prosthesis embodiment score was calculated using the average score from these five statements, taking the opposite (negative) value of the ‘foreign body’ statement.

### Statistical analysis

Statistical analysis was performed using IBM SPSS Statistics for Macintosh (Version 25) and JASP (Version 0.11.1). Tests for normality were carried out using a Shapiro-Wilk test, and statistical analysis was carried out using a repeated measures ANOVA for number of movements of each arm and non-parametric tests for MMR (Mann-Whitney). All correlations were performed using two-tailed Spearman correlation. An analysis of covariance (ANCOVA) with prosthesis use as a covariate was used to test the contribution of cause of limb-loss and type of prosthesis used. We further calculated the two-way random single measures of intraclass correlations (ICCs), allowing us to assess consistency of the PAL measurement. We also used a Kendall’s tau-b correlation to assess the consistency of wear-time frequency.

## Data Availability

All data will be available shortly at the Open Science Framework

## Acknowledgments

This work was supported by a Wellcome Trust Senior Research Fellowship (215575/Z/19/Z), and an ERC Starting Grant (715022 EmbodiedTech) and a Sir Henry Dale Fellowship jointly funded by the Wellcome Trust and the Royal Society (104128/Z/14/Z), awarded to T.R.M. M.MacS is funded by a Wellcome Trust Fellowship [100229/Z/12/Z]. We thank Harriet Dempsey Jones and Fiona van den Heiligenberg for developing the embodiment questionnaires and to Hunter Schone, Mischa Dhar and Victoria Root for data collection. We thank Opcare for help in participants recruitment, and our participants and their families for their ongoing support of our research.

---

**Table S1.** Demographic details of the participants involved in the study. Participant: AA = acquired amputee, AC = congenital one-hander, CO = two-handed control; participants marked with an asterisk were included in the gesticulation task. Y since amp = years since amputation. Gender: M = male, F = female. Amp Side = side of limb loss or non-dominant side: L = left, R = right. Amp level = level of limb loss: TR = trans-radial, TH = trans-humeral. Pros type = preferred type of prosthesis: Cos = cosmetic, Mech = mechanical, Myo = myo-electric. PLS = phantom limb sensation. PLP = phantom limb pain. SP = stump pain. Chronic PLS, PLP and SP were calculated by dividing maximum intensity of pain (0-100) by frequency (1 = all the time, 2 = daily, 3 = weekly, 4 = several times per month, and 5 = once or less per month). Pros Time = typical number of hours prosthesis worn per week. PAL = functional ability with prosthesis as determined by PAL questionnaire (0 = minimum function, 1 = maximum function). US = prosthesis usage score: +3 = maximum usage, -3 = minimum usage. EM = prosthesis embodiment score; +3 = maximum agreement with embodiment statements, -3 = maximum disagreement with embodiment statements.

| Participant | Age | Y Since Amp | Gender | Amp Side | Amp level | Amp cause | Prosthesis Type | PLS | PLP | SP | Pros wear time | PAL | US | EM |
| --- | --- | --- | --- | --- | --- | --- | --- | --- | --- | --- | --- | --- | --- | --- |
| AA01 | 58 | 14 | M | L | TR | Trauma | Myo | 15 | 0 | 0 | 119 | 0.5 | 1.87 | 2 |
| AA02 | 46 | 16 | F | L | TR | Trauma | Myo | 25 | 8 | 0 | 56 | 0.59 | 0.49 | -2.6 |
| AA03 | 50 | 3 | F | L | TR | Trauma | Mech | 0 | 0 | 0 | 77 | 0.44 | 0.44 | 2.4 |
| AA04 | 53 | 34 | M | L | TH | Trauma | Mech | 90 | 25 | 20 | 48 | 0.2 | -1.40 | 0 |
| AA05 | 21 | 1 | M | R | TR | Trauma | None | 50 | 30 | 17.5 | 0 | 0.04 | -3.44 | -3 |
| AA06* | 42 | 18 | M | R | TR | Trauma | Cos | 13.33 | 16 | 90 | 35 | 0.07 | -2.33 | -1.2 |
| AA07* | 61 | 21 | M | L | TR | Trauma | Cos | 95 | 50 | 0 | 105 | 0.67 | 2.21 | -2.2 |
| AA08* | 60 | 42 | M | R | TR | Trauma | Mech | 100 | 60 | 5 | 87.5 | 0.28 | 0.05 | -1.4 |
| AA09 | 65 | 37 | M | R | TH | Trauma | Mech | 90 | 0 | 12 | 98 | 0.46 | 1.11 | 3 |
| AA10 | 47 | 21 | M | R | TH | Trauma | Cos | 60 | 8 | 0 | 84 | 0.3 | 0.03 | 2 |
| AA11* | 68 | 12 | M | L | TR | Trauma | Mech | 0 | 0 | 7.5 | 35 | 0.54 | -0.31 | -1.8 |
| AA12* | 49 | 5 | M | R | TR | Vascular disease | Cos | 14 | 0 | 10 | 42 | 0.59 | 0.10 | 2 |
| AA13 | 57 | 29 | M | L | TR | Trauma | Mech | 6.25 | 0 | 17.5 | 65 | 0.11 | -1.31 | -0.2 |
| AA14* | 53 | 33 | M | L | TR | Trauma | Myo | 0 | 0 | 0 | 98 | 0.43 | 0.98 | 0.8 |
| AA15 | 28 | 10 | F | R | TR | Trauma | Mech | 85 | 21.67 | 0 | 2 | 0 | -3.55 | -2.8 |
| AA16 | 29 | 11 | M | L | TR | Trauma | None | 100 | 16 | 16 | 0 | 0 | -3.61 | -3 |
| AA17* | 43 | 20 | M | R | TR | Trauma | Myo | 37.5 | 0 | 10 | 98 | 0.61 | 1.75 | 2.4 |
| AA18* | 55 | 12 | M | L | TR | Trauma | Myo | 0 | 0 | 0 | 98 | 0.65 | 1.92 | -1.2 |
| AA19 | 61 | 17 | M | L | TR | Trauma | Mech | 30 | 17.5 | 20 | 91 | 0.74 | 2.11 | 1.4 |
| AA21* | 30 | 3 | M | L | TR | Trauma | Myo | 20 | 18 | 60 | 49 | 0.59 | 0.29 | 1.2 |
| AA22* | 46 | 5 | M | R | TR | Trauma | Myo | 70 | 2.5 | 16 | 56 | 0.57 | 0.40 | 1.8 |
| AC01 | 51 |  | F | L | TR | Congenital | Cos |  |  |  | 7 | 0.26 | -2.30 | -1.8 |
| AC02 | 47 |  | M | L | TR | Congenital | Mech |  |  |  | 84 | 0.7 | 1.75 | 0.2 |
| AC03* | 45 |  | F | L | TR | Congenital | Myo |  |  |  | 63 | 0.46 | 0.13 | -0.4 |
| AC04* | 26 |  | M | L | TR | Congenital | Mech |  |  |  | 6 | 0.13 | -2.88 | -1.4 |
| AC05* | 55 |  | F | L | TR | Congenital | Cos |  |  |  | 112 | 0.3 | 0.82 | -0.8 |
| AC06* | 63 |  | M | L | TR | Congenital | Cos |  |  |  | 87.5 | 0.35 | 0.35 | 2.2 |
| AC07 | 35 |  | M | L | TR | Congenital | Cos |  |  |  | 56 | 0.28 | -0.84 | 1.6 |
| AC08* | 26 |  | F | L | TR | Congenital | Cos |  |  |  | 84 | 0.24 | -0.22 | -0.4 |
| AC09* | 49 |  | M | L | TR | Congenital | Myo |  |  |  | 91 | 0.57 | 1.39 | 1.8 |
| AC10 | 42 |  | M | L | TR | Congenital | Cos |  |  |  | 56 | 0.54 | 0.28 | -1 |
| AC11 | 66 |  | F | R | TR | Congenital | Cos |  |  |  | 42 | 0.35 | -0.93 | 0 |
| AC12* | 56 |  | F | R | TR | Congenital | Cos |  |  |  | 98 | 0.43 | 0.98 | 2 |
| AC13 | 53 |  | M | L | TH | Congenital | Mech |  |  |  | 63 | 0.33 | -0.43 | 2.4 |
| AC14 | 42 |  | M | L | TR | Congenital | Mech |  |  |  | 2 | 0.09 | -3.17 | -0.6 |
| AC15* | 55 |  | F | L | TR | Congenital | Myo |  |  |  | 105 | 0.65 | 2.12 | 2 |
| AC16* | 38 |  | F | R | TR | Congenital | Cos |  |  |  | 84 | 0.67 | 1.62 | 2 |
| AC17* | 29 |  | M | L | TR | Congenital | Myo |  |  |  | 70 | 0.46 | 0.33 | 0.4 |
| AC18* | 53 |  | F | L | TR | Congenital | Cos |  |  |  | 48 | 0.65 | 0.52 | 3 |
| AC20* | 52 |  | F | R | TR | Congenital | Myo |  |  |  | 32.5 | 0.26 | -1.58 | 0 |
| AC21* | 32 |  | F | R | TR | Congenital | Myo |  |  |  | 40 | 0.41 | -0.73 | 2 |
| AC22 | 57 |  | M | R | TR | Congenital | Mech |  |  |  | 126 | 0.69 | 2.88 | 1.8 |
| AC23* | 47 |  | F | L | TR | Congenital | Myo |  |  |  | 84 | 0.89 | 2.56 | 3 |
| AC25* | 41 |  | M | L | TR | Congenital | Myo |  |  |  | 112 | 0.85 | 3.17 | 3 |
| CO01* | 48 |  | M |  |  |  |  |  |  |  |  |  |  |  |
| CO04* | 59 |  | M |  |  |  |  |  |  |  |  |  |  |  |
| CO05* | 27 |  | F |  |  |  |  |  |  |  |  |  |  |  |
| CO07* | 35 |  | M |  |  |  |  |  |  |  |  |  |  |  |
| CO08* | 34 |  | F |  |  |  |  |  |  |  |  |  |  |  |
| CO10* | 70 |  | M |  |  |  |  |  |  |  |  |  |  |  |
| CO12* | 18 |  | F |  |  |  |  |  |  |  |  |  |  |  |
| CO13* | 67 |  | M |  |  |  |  |  |  |  |  |  |  |  |
| CO14* | 50 |  | M |  |  |  |  |  |  |  |  |  |  |  |
| CO15* | 51 |  | F |  |  |  |  |  |  |  |  |  |  |  |
| CO16* | 36 |  | F |  |  |  |  |  |  |  |  |  |  |  |
| CO17* | 41 |  | M |  |  |  |  |  |  |  |  |  |  |  |
| CO18* | 33 |  | M |  |  |  |  |  |  |  |  |  |  |  |
| CO19* | 45 |  | M |  |  |  |  |  |  |  |  |  |  |  |
| CO21* | 54 |  | M |  |  |  |  |  |  |  |  |  |  |  |

**Supplementary Figure 1.**
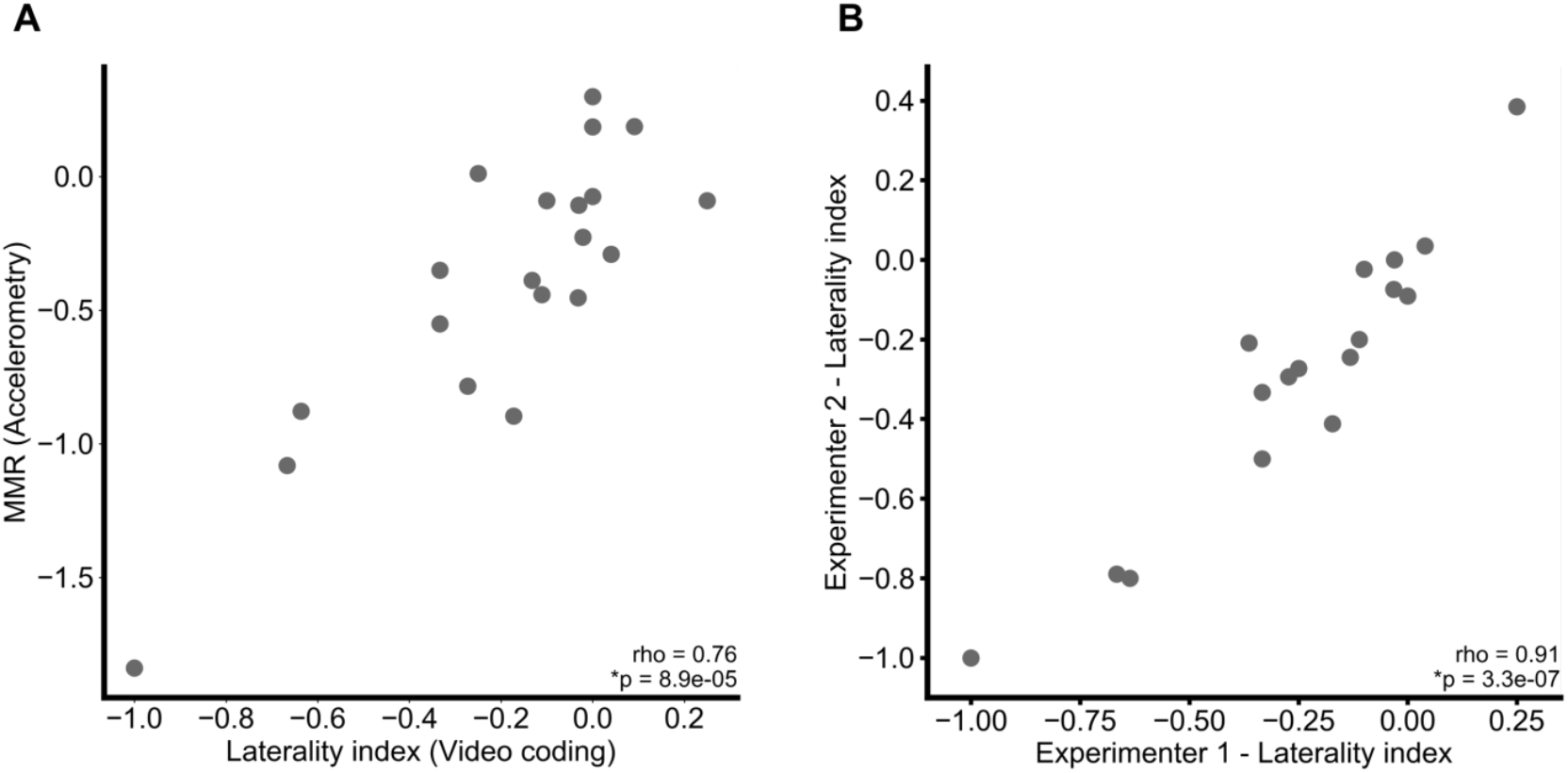
Measurement validation using Offline video coding. (A) Correlation between the laterality index calculated from offline video coding, on a subset of participants, and the MMR (rho_(18)_ = 0.76, p <0.001). (B) Test-retest reliability of the offline video coding method. The laterality measure was validated by an additional experimenter. The measurement was found to be stable across the two separate experimenters (rho_(16)_ = 0.96, p < 0.001).

**Supplementary Figure 2.**
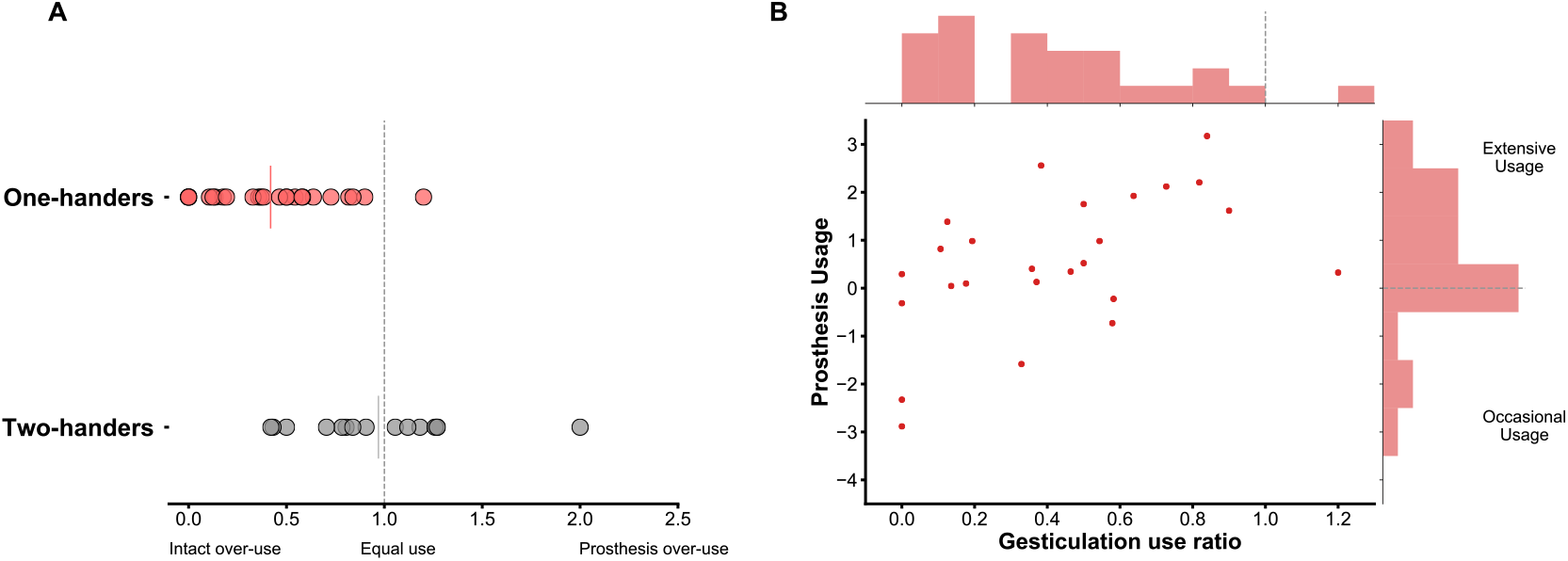
Results of analysis with alternative gesture measure: Use ratio. (A) Use ratio across groups; two-handers performed movements with both hands equally when gesturing, while one-handers were significantly lateralised towards their intact hand (U_(38)_ = 54, p < 0.001). Solid coloured vertical lines indicate the group mean (B) Increased daily usage associated with increased incorporation of the prosthesis into gestures as measured by the use ratio (rho_(23)_ = 0.55, p = 0.004).

**Supplementary Figure 3.**
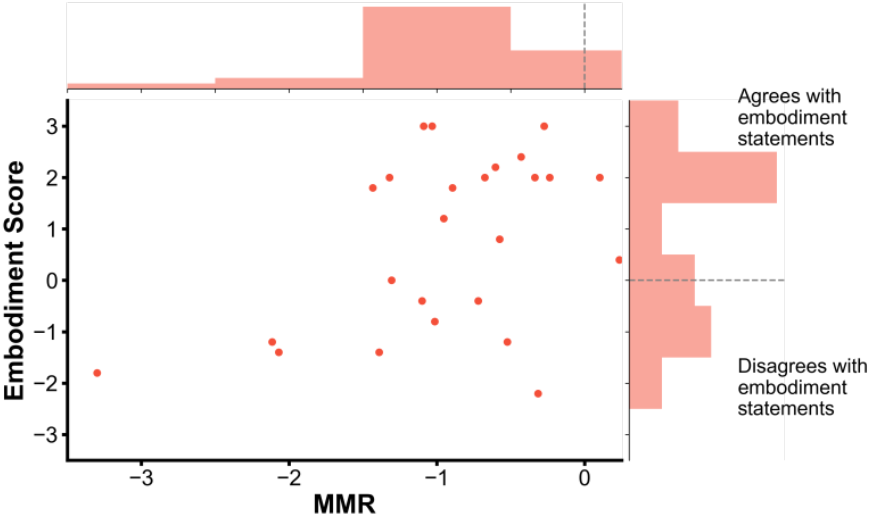
Embodiment and gesture behaviour. Greater perceived embodiment was loosely associated with increased incorporation of the prosthesis into gestures as measured by MMR (rho_(23)_ = 0.37, p = 0.07).

## References

Alibali, M. W., Heath, D. C., & Myers, H. J. (2001). Effects of Visibility between Speaker and Listener on Gesture Production: Some Gestures Are Meant to Be Seen. Journal of Memory and Language, 44(2), 169–188. https://doi.org/10.1006/jmla.2000.2752

Bailey, R. R., Klaesner, J. W., & Lang, C. E. (2015). Quantifying Real-World Upper-Limb Activity in Nondisabled Adults and Adults With Chronic Stroke. Neurorehabilitation and Neural Repair, 29(10), 969–978. https://doi.org/10.1177/1545968315583720

Chadwell, A., Kenney, L., Thies, S., Galpin, A., & Head, J. (2016). The reality of myoelectric prostheses: Understanding what makes these devices difficult for some users to control. Frontiers in Neurorobotics, 10(AUG). https://doi.org/10.3389/fnbot.2016.00007

Engdahl, S. M., Christie, B. P., Kelly, B., Davis, A., Chestek, C. A., & Gates, D. H. (2015). Surveying the interest of individuals with upper limb loss in novel prosthetic control techniques. Journal of Neuroengineering and Rehabilitation, 12(1), 53. https://doi.org/10.1186/s12984-015-0044-2

Feyersein, P., & De Lannoy, J. D. (1991). Gestures and Speech: psychological investigation. New York: Cambridge University Press.

Goldin-Meadow, S. (2003). Hearing gesture: How our hands help us think. Cambridge, MA.: Harvard University Press.

Goldin-Meadow, S., & Alibali, M. W. (2013). Gesture’s Role in Speaking, Learning, and Creating Language. Annual Review of Psychology, 64(1), 257–283. https://doi.org/10.1146/annurev-psych-113011-143802

Iverson, J. M., & Goldin-Meadow, S. (1998). Why people gesture when they speak. Nature, 396(6708), 228. https://doi.org/10.1038/24300

Jang, C. H., Yang, H. S., Yang, H. E., Lee, S. Y., Kwon, J. W., Yun, B. D., … Jeong, H. W. (2011). A Survey on Activities of Daily Living and Occupations of Upper Extremity Amputees. Annals of Rehabilitation Medicine, 35, 907. https://doi.org/10.5535/arm.2011.35.6.907

Lang, C. E., Waddell, K. J., Klaesner, J. W., & Bland, M. D. (2017). A Method for Quantifying Upper Limb Performance in Daily Life Using Accelerometers. Journal of Visualized Experiments, (122), 1–8. https://doi.org/10.3791/55673

Lausberg, H., & Slöetjes, H. (2008). Gesture coding with the NGCS - ELAN system. Proceedings of Measuring Behavior 2008, 2008, 176–177.

Longo, M. R., Schüür, F., Kammers, M. P. M., Tsakiris, M., & Haggard, P. (2008). What is embodiment? A psychometric approach. Cognition, 107(3), 978–998. https://doi.org/10.1016/j.cognition.2007.12.004

Lu, J. C., & Goldin-Meadow, S. (2018). Creating Images With the Stroke of a Hand: Depiction of Size and Shape in Sign Language. Frontiers in Psychology, 9(July). https://doi.org/10.3389/fpsyg.2018.01276

Maimon-Mor, R. O., & Makin, T. R. (2020). Is an artificial limb embodied as a hand? Brain decoding in prosthetic limb users. Under Review.

Makin, T. R., Cramer, A. O., Scholz, J., Hahamy, A., Henderson Slater, D., Tracey, I., & Johansen-Berg, H. (2013). Deprivation-related and use-dependent plasticity go hand in hand. ELife, 2013, 1–15. https://doi.org/10.7554/eLife.01273.01273

Makin, T. R., de Vignemont, F., & Faisal, A. A. (2017). Neurocognitive barriers to the embodiment of technology. Nature Biomedical Engineering, 1(1), 0014. https://doi.org/10.1038/s41551-016-0014

McNeill, D. (1992). Hand and mind: What gestures reveal about thought. Chicago and London: The University of Chicago Press.

McNeill, D., & Levy, E. T. (1982). Conceptual Representations in Language Activity and Gesture. In R. J. Jarvella & W. Klein (Eds.), Speech, Place, and Action (pp. 271–295). Wiley.

Østlie, K., Lesjø, I. M., Franklin, R. J., Garfelt, B., Skjeldal, O. H., & Magnus, P. (2012). Prosthesis rejection in acquired major upper-limb amputees: a population-based survey. Disability and Rehabilitation: Assistive Technology, 7(4), 294–303. https://doi.org/10.3109/17483107.2011.635405

Pareés, I., Saifee, T. A., Kassavetis, P., Kojovic, M., Rubio-Agusti, I., Rothwell, J. C., … Edwards, M. J. (2012). Believing is perceiving: Mismatch between self-report and actigraphy in psychogenic tremor. Brain, 135(1), 117–123. https://doi.org/10.1093/brain/awr292

Uswatte, G., Taub, E., Morris, D., Light, K., & Thompson, P. A. (2006). The Motor Activity Log-28: assessing daily use of the hemiparetic arm after stroke. Neurology, 67(7), 1189–1194. https://doi.org/10.1212/01.wnl.0000238164.90657.c2

Van Den Heiligenberg, F. M. Z., Orlov, T., MacDonald, S. N., Duff, E. P., Henderson Slater, D., Beckmann, C. F., … Makin, T. R. (2018). Artificial limb representation in amputees. Brain, 141(5), 1422–1433. https://doi.org/10.1093/brain/awy054

van den Heiligenberg, F. M. Z., Yeung, N., Brugger, P., Culham, J. C., & Makin, T. R. (2017). Adaptable Categorization of Hands and Tools in Prosthesis Users. Psychological Science, 28(3), 395–398. https://doi.org/10.1177/0956797616685869

Vujaklija, I., Farina, D., & Aszmann, O. C. (2016). New developments in prosthetic arm systems. Orthopedic Research and Reviews, 8, 31–39. https://doi.org/10.2147/ORR.S71468

